# An AI-Assisted Study on EEG Spectral Power Gradients in Children with Monogenic Epilepsy

**DOI:** 10.64898/2026.09.17.26363339

**Authors:** Alan Liang, Wim van Drongelen, Douglas R. Nordli

## Abstract

Based on the observation that in normal EEG rhythms, the low-voltage beta frequencies dominate in the anterior region while characteristic high(er)-amplitude slower rhythms manifest more posteriorly, we studied these gradients across normal subjects and patients with different monogenic epilepsies. From large data sets (long term records of 374 controls and 241 patients) we selected 60s eyes-open rest-EEG epochs to determine the spectral power of the EEG bands. To help manage the large data sets and facilitate analyses, we employed AI to preselect and assist in coding. While both preselection and coding was extensively verified and validated by the authors, which was a time-consuming effort, the AI contribution reduced the net effort of evaluating the large dataset.

We developed a metric that reflects the anterior-posterior gradients of the beta rhythm and the so-called posterior dominant rhythm. Our results indicate that this simple gradient measure across 1-10 year-age groups may provide a robust quantitative biomarker of cerebral dysfunction in monogenic epilepsy. Because EEG is noninvasive, widely available, repeatable, and relatively inexpensive, such a biomarker could ultimately provide a practical means of assessing the cerebral effects of emerging targeted therapies across multiple genetic epilepsies.

## INTRODUCTION

Electroencephalography (EEG) remains an important tool for diagnosing, classifying, and managing epilepsy. Capturing a live seizure provide important diagnostic information, but seizures occur unpredictably and often infrequently. Therefore, ictal monitoring may require prolonged recording, which makes it highly time-consuming, expensive, and stressful for patients. Such limitation motivates the research for interictal EEG biomarkers that can support disease assessment without requiring seizure capture (e.g., Gallotto and Seeck 2023; Staba et al., 2014; Kamintsky et al., 2023; Gnatkovsky et al., 2026). The need for quantitative interictal EEG biomarker is equally relevant to monogenic epilepsies. These disorders result from mutation in single gene, demonstrating diverse clinical phenotypes and neurodevelopmental impairment (Guerrini et al., 2021). This clinical heterogeneity motivates the search for interictal EEG biomarkers that can identify the shared underlying abnormality to facilitate disease diagnosis and evaluation of treatment.

Interictal EEG contains information for both epileptiform discharges (IEDs) and background EEG rhythms. Background EEG activity comprises ongoing electrical activity that varies with frequency, amplitude, spatial distribution, and organization. The advent of digital signal processing and machine learning enables the quantitative analysis of various properties derived from the background EEG rhythm, extending the EEG biomarker landscape to both observed properties of the EEG and those sub-visual features.

Previous studies have identified quantitative abnormalities in background EEG rhythms across several types of epilepsy. This paragraph lists a few examples. In children with SCN1A-related Dravet syndrome, spectral analysis on resting state-EEG revealed decrease in relative alpha power and increase in relative theta power with respect to age compared to healthy controls (Holmes et al., 2012). In STXBP1-related epilepsy, increased relative delta power showed an anterior predominance and was associated with higher clinical severity (Cossu et al., 2024). More recently, regional spectral features have been used to distinguish patients with monogenic epilepsies from controls and associate with neurological outcome (Galer et al., 2025). Together, these studies motivate further spectral analysis of background EEG as potential biomarker, its developmental trend, and its association with clinical severity.

In addition to perform spectral analysis across scalp or within predefined brain regions, we examined the topographic organization of power of EEG rhythms along the anterior-posterior axis during brain development. Our investigation is based on the observation that in normal EEG rhythms, the low-voltage faster frequencies dominate in the anterior region while high(er)-amplitude slower rhythms manifest more posteriorly. Therefore, we evaluated the gradients of these rhythms—the beta rhythm and posterior dominant rhythm—across age groups and tested their suitability as a biomarker for different types of monogenic epilepsy.

A short description of this work was submitted as an Abstract for the AES 2026 meeting.

## METHODS

### Normal Subjects and Patients

De-identified data from healthy controls were obtained from the Brain Data Science Platform (Zafar, et al., 2025), and patient recordings were obtained from Ciitizen Health (https://www.citizen.health). The work presented here was approved by the University of Chicago IRB. Our analysis included 374 controls and 241 patients with different types of monogenic epilepsy (KCNT1, FOXG1, WWOX, SCN8A, PCDH19, SCN2A, SYNGAP1) (Table 1).

**Table 1.**
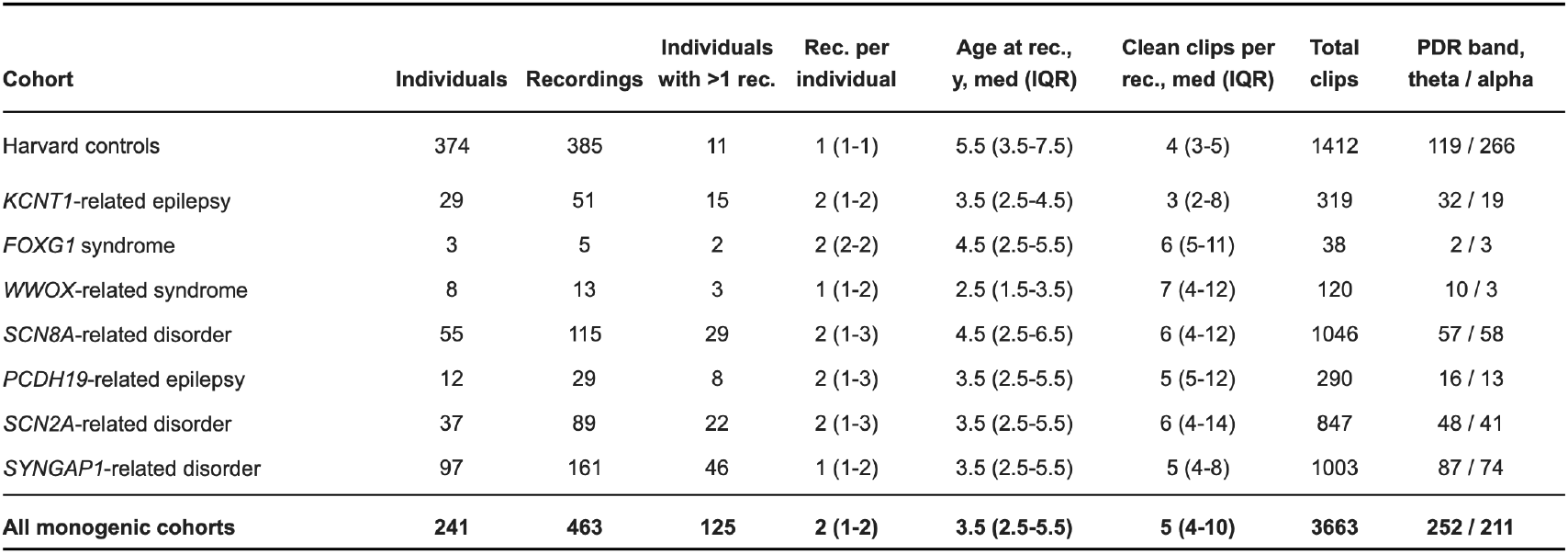
Overview of the Dataset.

| Cohort | Individuals | Recordings | Individuals with >1 rec. | Rec. per individual | Age at rec., y, med (IQR) | Clean clips per rec., med (IQR) | Total clips | PDR band, theta / alpha |
| --- | --- | --- | --- | --- | --- | --- | --- | --- |
| Harvard controls | 374 | 385 | 11 | 1 (1-1) | 5.5 (3.5-7.5) | 4 (3-5) | 1412 | 119 / 266 |
| KCNT1-related epilepsy | 29 | 51 | 15 | 2 (1-2) | 3.5 (2.5-4.5) | 3 (2-8) | 319 | 32 / 19 |
| FOXG1 syndrome | 3 | 5 | 2 | 2 (2-2) | 4.5 (2.5-5.5) | 6 (5-11) | 38 | 2 / 3 |
| WWOX-related syndrome | 8 | 13 | 3 | 1 (1-2) | 2.5 (1.5-3.5) | 7 (4-12) | 120 | 10 / 3 |
| SCN8A-related disorder | 55 | 115 | 29 | 2 (1-3) | 4.5 (2.5-6.5) | 6 (4-12) | 1046 | 57 / 58 |
| PCDH19-related epilepsy | 12 | 29 | 8 | 2 (1-3) | 3.5 (2.5-5.5) | 5 (5-12) | 290 | 16 / 13 |
| SCN2A-related disorder | 37 | 89 | 22 | 2 (1-3) | 3.5 (2.5-5.5) | 6 (4-14) | 847 | 48 / 41 |
| SYNGAP1-related disorder | 97 | 161 | 46 | 1 (1-2) | 3.5 (2.5-5.5) | 5 (4-8) | 1003 | 87 / 74 |
| All monogenic cohorts | 241 | 463 | 125 | 2 (1-2) | 3.5 (2.5-5.5) | 5 (4-10) | 3663 | 252 / 211 |

### Use of AI

We employed AI (ChatGPT) to assist with EEG record selection, analysis procedures and parts of the text. In all cases, the authors evaluated results.

In the <u>EEG selection</u>, the final selection was reviewed by the pediatric electroencephalographer (DRN) of our team. Details are further described in the next Section on automated identification.

<u>Analyses procedures</u> were evaluated by all of us. The AI-software generation and evaluation consisted of (1) providing the metrics (see Section on Spectral Power Gradients), (2) challenging the algorithms subsequently proposed by the AI application—several challenges and iterations were required to obtain a correct procedure—then, (3) using test signals with known spectral content to validate the final algorithms, and (4) verifying the spectral outcomes of the AI software on a subgroup with home-made Matlab scripts written by AL and WvD. After AI-software passed all three validation steps, we used it for further statistical analyses.

For a parts of the text we <u>used AI to write a draft.</u> These drafts were then edited by all of us. Most drafts were well-written, but none passed without serious editing by the authors.

### Automated identification and screening of awake EEG segment

To facilitate quantitative analysis across large EEG datasets, we developed a semi-automated Python procedure (version 3.10.12) to identify candidate segments of artifact-free awake EEG. Because the duration and behavioral composition of the source recordings varied substantially, the objective was not to analyze the complete EEG but rather to identify representative 1–2-minute epochs of wakefulness that could subsequently undergo standardized quantitative analysis.

Candidate awake segments were identified using physiologic and signal-quality features intended to distinguish wakefulness from sleep and artifact. Particular emphasis was placed on the presence of spontaneous eye blinks, which provided a useful marker of behavioral wakefulness in recordings for which synchronized video or detailed behavioral annotation was not consistently available. Candidate segments were screened for an appropriate rate and spatial distribution of blink transients, with frontal predominance used to distinguish probable eye movements from generalized EEG transients. The algorithm additionally evaluated signal quality and rejected or penalized intervals containing excessive artifact, prolonged low-amplitude or flat signals, or other features incompatible with reliable quantitative analysis. Sleep-related activity, particularly the presence of spindle-range activity, was used as evidence against wakefulness. When developmentally appropriate, the presence and frequency of posterior rhythmic activity were also considered in identifying plausible awake segments.

Rather than treating automated detection as a substitute for expert EEG interpretation, the algorithm was designed as a candidate-generation and screening tool. For each recording, candidate segments were exported for visual review, together with EEG displays that allowed rapid assessment of state and signal quality. Candidate epochs were then reviewed by a pediatric electroencephalographer (DRN). Segments containing sleep, excessive movement or electrode artifact, ictal activity, or other features likely to confound quantitative measurements were excluded. This human-in-the-loop approach substantially reduced the amount of EEG requiring manual review while retaining expert confirmation of behavioral state and data quality. Importantly, the same general selection framework was used for the normative and disease cohorts, thereby reducing the potential for systematic differences in epoch selection between groups.

### Spectral Power Gradients

The resulting curated awake segments were subsequently processed using a common preprocessing and quantitative-analysis pipeline. Analyses were performed on standardized bipolar derivations spanning the anterior-to-posterior axis, permitting direct comparison of spatial and spectral EEG organization between children with monogenic epilepsies and age-matched normative controls. Representative samples are depicted in Figure 1.

**Figure 1.**
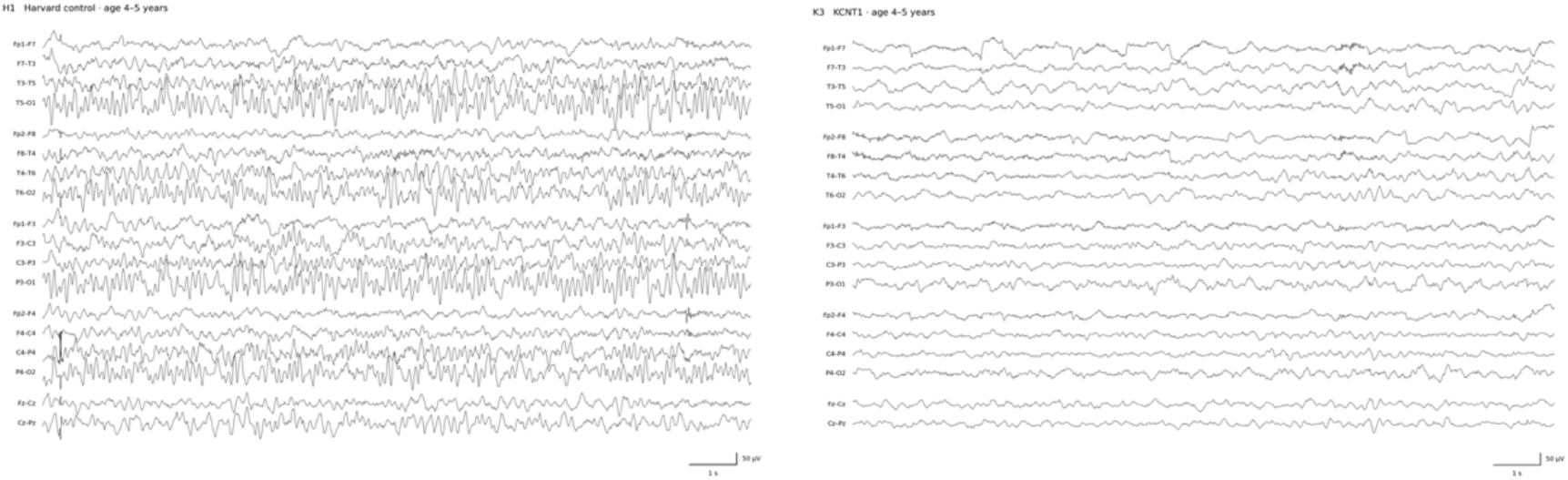
Two representative EEG Examples from a control subject (left panel) and a monogenic patient with a KCNT1 mutation (right panel).

From each selected 60s rest-EEG epoch (bandwidth: 1-70Hz) we determined the power of the EEG bands: Delta (1.5-4.0Hz), Theta (4.0-8.0Hz), Alpha (8.0-13.0Hz) and Beta (13.0-20.0Hz). The posterior dominant rhythm metric we employed was the power in the theta band for subjects/patients <4 years old and the power in the alpha band for the older group. We used filters to obtain the signal within each of the above frequency bands, and the Teager-Kaiser Operator (TKO) to obtain a power metric (see Appendix for details, and Kaiser, 1990). We used several montages to asses robustness of topographic properties of the EEG rhythms, the data we show in Figures 2 and 3 was obtained with Fp1-F3, F3-C3, C3-P3, P3-O1, Fp2-F4, F4-C4, C4-P4, P4-O2.

**Figure 2.**
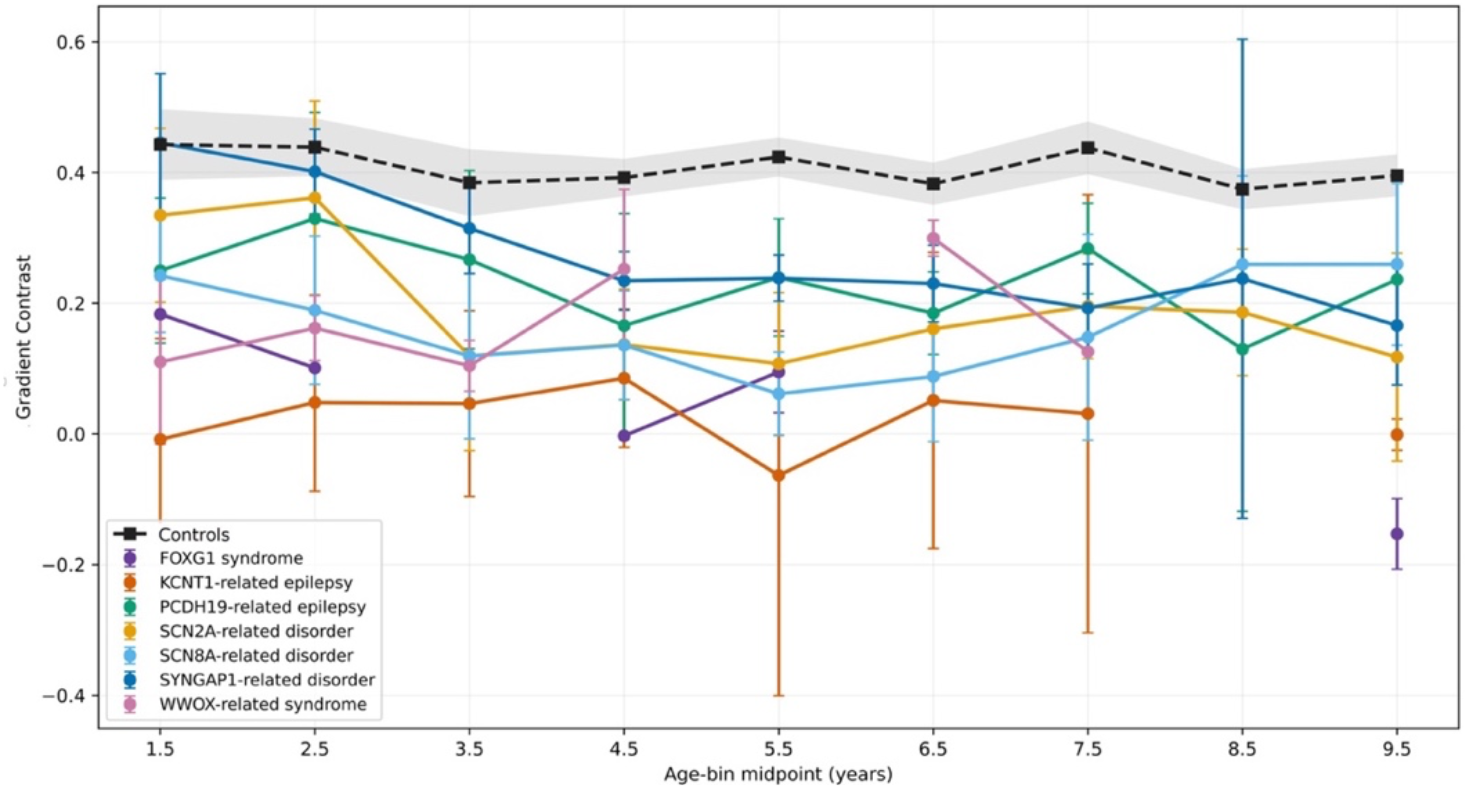
Overview of the Age-matched TKO-based Gradient Contrast for controls and patient groups with monogenic epilepsy. Confidence intervals (95%) are indicated by the vertical bars (patients) and the grey shaded area (controls). The initial age-bins of patients with the SYNGAP-1 and the SCN2A mutations are close to the normal values.

**Figure 3.**
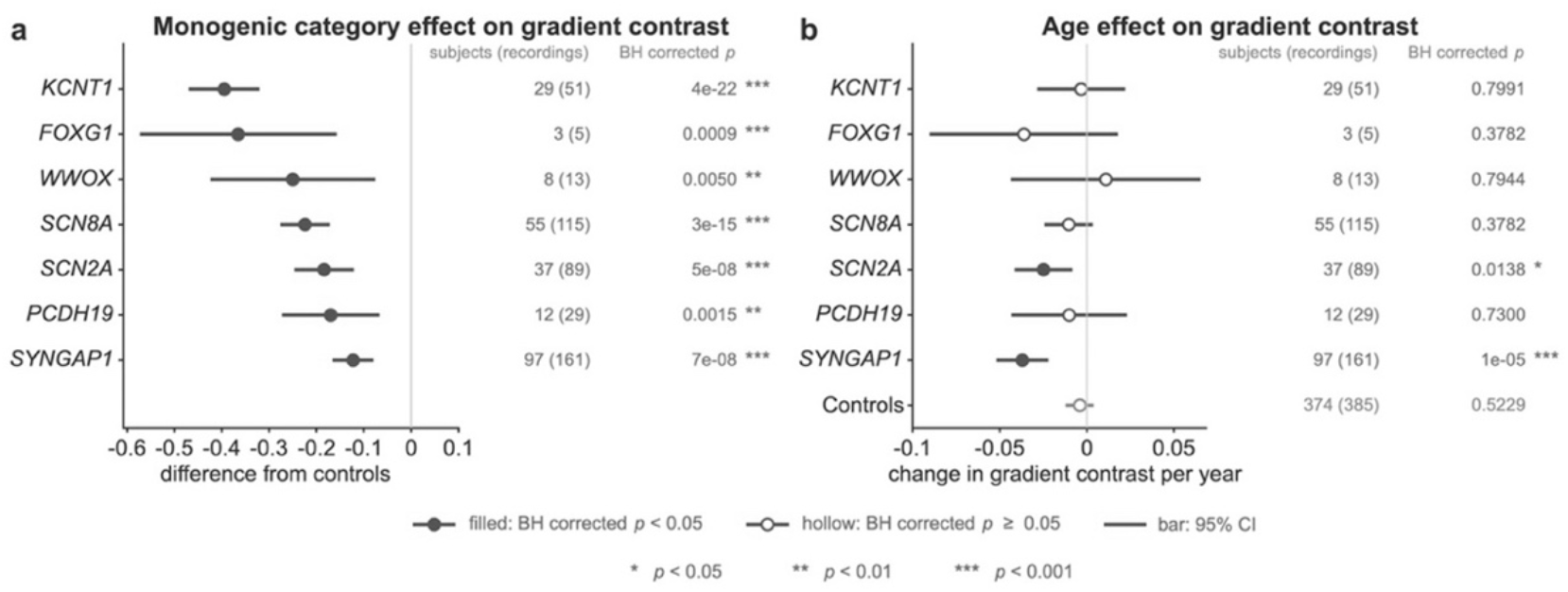
Linear Mixed Model analysis of the results shown in Fig. 2. (a) Metrics in the groups with a mutation were significantly different from the normals. (b) Metrics were not significantly different across the age groups, except in the patients with the SYNGAP-1 and the SCN2A mutations. BH corrected = Benjamini-Hochberg correction for multiple testing.

Next we applied the following computational steps.

<u>Step 1</u>: We computed the power metrics. Delta (Pd, 1.5-4.0Hz), theta (Pt, 4.0-8.0Hz), alpha (Pa, 8.0-13.0Hz) and beta (Pb, 13.0-20.0Hz)

<u>Step2</u>: For each frequency band we computed the sum of these metrics for the left channels (Fp1-F3, F3-C3, C3-P3, P3-O1) and the sum of the right channels (Fp2-F4, F4-C4, C4-P4, P4-O2).

<u>Step 3</u>: The left and right hemisphere sums were used to normalize the individual band-powers: e.g., for the alpha rhythm in the F4-C4 signal we computed [Pa(F4-C4)]/[SumAlpha(Right)], for the theta rhythm in C3-P3 we computed [Pt(C3-P3)]/[SumTheta(Left)], etc.

<u>Step 4</u>: For frequency bands we computed the anterior-posterior gradients (G) of the normalized metrics. To avoid montage related correlation, we made sure that no common electrodes were present in the differences, e.g., G(Left, Theta)= [Pt(Fp1-F3)]/[SumTheta(Left)]-[Pt(P3-O1)]/[SumTheta(Left)], G(Right, Alpha)= [Pa(Fp2-F4)]/[SumAlpha(Right)]-[Pa(P4-O2)]/[SumAlpha(Right)], etc.

<u>Step 5</u>: We determined left and right Gradient Contrasts (GC) as the difference between the beta gradient and the gradient of the band of the posterior dominant rhythm. For the estimate of the posterior dominant rhythm, we made an age distinction: below 4 years of age the theta band metrics were used and above 4 years we employed the alpha band values. Some examples: for a 2 year old, GC(Right)= G(Right,Beta)-G(Right,Theta) and for a 5 year old, GC(Left)= G(Left,Beta)-G(Left,Alpha), etc.

<u>Step 6</u>: The left and right results for step 5 were averaged in a Gradient Contrast GC(Average), that was used in the subsequent statistical analyses and e.g., the results depicted in Figs. 2 and 3.

### Statistics

Statistics analyses were performed in MATLAB 2025b using the Statistics and Machine Learning Toolbox (The MathWorks, Inc., Natick, Massachusetts, United States) and R (Version 4.4.1) for Receiver Operator Characteristic (ROC) analyses. The analysis includes 848 recordings from 615 individuals, covering one-year age bins from 1 to 10 years. Age for each recording was represented by the midpoint of each bin (e.g. 1.5, 2.5…9.5 years) (see Appendix for details). We fitted the effects of mutation and age as fixed effect covariate on the GC metrics derived from each recording using a linear mixed effect model, where subject-specific random intercept is modeled to account for the correlation among repeated observation. The model was estimated using restricted maximum likelihood method (see Appendix for details).

We first tested whether the GC for patient is statistically different from control. Furthermore, to assess the age effect on GC for each control and monogenic mutation groups, we tested if the estimated age slope differed from zero in the linear mixed model. Hypothesis were evaluated using two-sided t-tests, with degrees of freedom for these tests estimated by Satterthwaite approximation. Multiple hypothesis testing in the result was corrected by Benjamini-Hochberg(BH) procedure (see Appendix for details).

To ensure the stability of the model parameter estimation and hypothesis testing, we visually verified the assumption for our statistical model: the distribution of residues and random effect were approximately normal and homoscedastic using the Q-Q plot and residual-versus-fit plot, respectively.

The capability of using the gradient contrast as biomarker to distinguish patients and control is assessed using the clustered-based ROC method to avoid repeated recording under or overestimated the efficacy of ROC (see Appendix for details).

## RESULTS

Two representative awake rest-EEG records used for our analysis are shown in Figure 1. The result of the Teager-Kaiser Operator (TKO) applied to the data, summarized in Table 1, are depicted in Figures 2-4 and Table 2.

**Table 2.** Summary of the clustered ROC result—AUROC values and 95% confidence intervals—per mutation and for two age groups.

| Genetic group | Age 1-4 years |  |  | Age 4-10 years |  |  |
| --- | --- | --- | --- | --- | --- | --- |
|  | Individuals | Recordings | AUROC (95% CI) | Individuals | Recordings | AUROC (95% CI) |
| *KCNT1* | 23 | 32 | 0.858 (0.772-0.945) | 14 | 19 | 0.946 (0.870-1.000) |
| *FOXG1* | 2 | 2 | 0.849 | 2 | 3 | 0.994 |
| *WWOX* | 7 | 10 | 0.849 (0.777-0.921) | 2 | 3 | 0.812 |
| *SCN8A* | 40 | 57 | 0.743 (0.657-0.829) | 33 | 58 | 0.850 (0.785-0.915) |
| *PCDH19* | 9 | 16 | 0.680 (0.565-0.795) | 9 | 13 | 0.816 (0.739-0.892) |
| *SCN2A* | 31 | 48 | 0.637 (0.528-0.747) | 19 | 41 | 0.890 (0.849-0.930) |
| *SYNGAP1* | 63 | 87 | 0.553 (0.467-0.640) | 52 | 74 | 0.803 (0.749-0.858) |

The mean gradient contrast (GC(Average)) was plotted against age at the time of recordings for controls and all monogenic epilepsy cohorts.

Statistical analysis confirmed the observation in Figure 2, i.e., that controls had significantly higher gradient contrast than the monogenic epilepsy cohort. The outcomes for the data in Figure 2 are summarized in Figure 3a. In addition, analysis demonstrated that, except for SYNGAP1 and SCN2A, gradient contrast was not significantly related to age (Fig. 3b).

We performed Receiver Operator Characteristic (ROC) analysis to further assess the value of the GC as a biomarker. We determined the Area Under the Receiver Operating Characteristic curve (AUROC) for the each of the patient cohorts relative to the normal group for the 1-4 years old and the 4-10 years old cohorts (Fig. 4). Comparing the panels in Figure 4, it can be seen that overall, the AUROC scores higher in the older group. Also, in agreement with the results presented in Figure 3, the ranges in the left panel of Figure 4 show that KCNT1, FOXG1, and WWOX show a greater difference versus the normal group and that SYNGAP1, SCN2A, PCDH19, and SCN8A land in a category with a smaller AUROC score.

**Figure 4.**
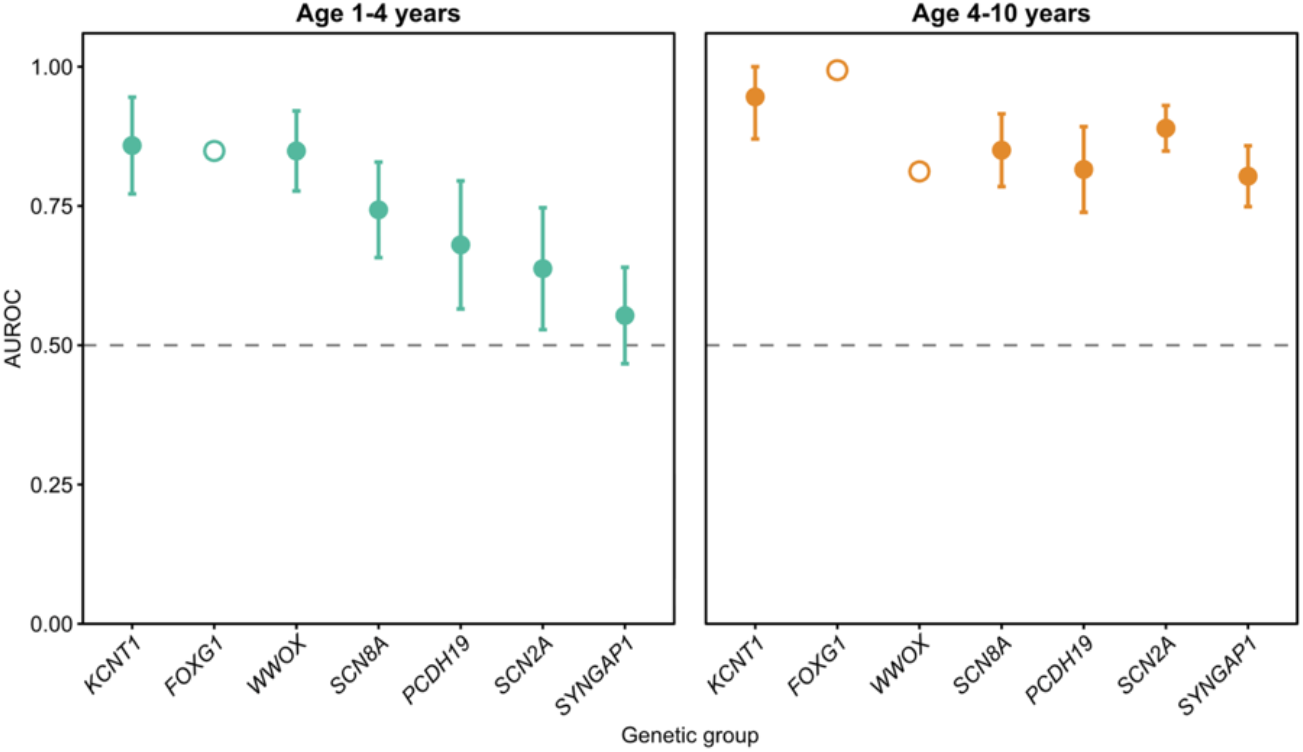
Clustered ROC analysis of gradient contrast. Point represent AUROC estimate, and the error bar represent the 95% confidence interval. Open circles identify the comparisons with fewer than five patient subject, where confidence intervals were not included due to limited sample size.

The details of the AUROC analysis, values per mutation and associated confidence intervals are summarized in Table 2.

## DISCUSSION

We found that the gradient contrast derived from rest-EEG can be used to distinguish between controls and patients with epilepsy (Figs. 2-4). In addition, we observed differences in the gradient contrast values between the different disorders and, in some cases, an apparent progression within disease cohorts (SYNGAP1 and SCN2A) (Figs. 3, 4). In sum, our results with the anterior-posterior gradients of the beta rhythm and the posterior dominant rhythm indicate that this metric across 1-10 year-age groups shows potential as a robust quantitative biomarker of cerebral dysfunction in monogenic epilepsy. Because EEG is noninvasive, widely available, repeatable, and relatively inexpensive, such a biomarker could ultimately provide a practical means of assessing the cerebral effects of emerging targeted therapies across multiple genetic epilepsies.

As a general rule, the validity of signal processing results depends on the combination of all procedures that are applied—from recording to final analyses—plus the quality and quantity of the input data. Analysis of the electroencephalogram (EEG) is no exception to this rule. And, because EEG is very susceptible to artifact—i.e., to be contaminated with biological or non-biological signals originating from outside the brain—gathering good input can be a challenge for quantitative analyses of EEG (qEEG). While the experienced clinical electrophysiologist reads around EEG artifacts, qEEG procedures mostly rely on automated artifact rejection. The automated rejection algorithms are not without caveats: e.g., rejection of large amplitude deflections may result in the undesired removal of discharges from brain networks, and the use of independent component analysis (ICA) —popular for removing eye artifact—may result in undesired removal of frontal delta rhythms. In the current digital-world, artificial intelligence (AI) starts to play a role in these types of signal analysis as well. A compelling recent publication describes the development of automated EEG interpretation (Sun et al., 2026). While this is a great, useful addition to the tools available to the EEG-world, AI because of its black-box characteristic is not without potential problems. Misunderstandings between the users and AI application can lead to misinterpretation of the AI generated outcomes. For this reason we employed a semi-automatic procedure where automated procedures generated EEG epoch candidates that were subjected to a final reviewed by a clinical electroencephalographer. Furthermore, analysis algorithms proposed by AI were often incorrect and several iterations were required to produce the correct underlying mathematics and test results. In sum, we found that challenging any AI application as well as testing and validating input-output relationships of its procedures are time-consuming and essential for automated research applications. Nonetheless, in our experience, the application of AI saves significant amounts of time, and we found that it is useful and efficient in the type of project presented here.

## Data Availability

EEG data are available at the sources listed in the Reference section.
All further data produced in the present study are available upon reasonable request to the authors

## Appendix

### Details on the Applied Power Metric and Statistical Models

#### The Teager-Kaiser Operator

Neuronal propagation lags are bound by conduction and transmission. As a consequence, the lower frequency components are subjected to less of a phase shift as compared to the higher frequency ones. Therefore, it is likely that the power of low frequency local field potentials (LFPs) will cancel each other less than those of the higher frequencies. Hence, one might expect that the power in the LFP and EEG signals decrease with frequency and therefore the power will not be a good reflection of the activity levels associated with the generators of these rhythms. As will be outlined in the following, the TKO approach and/or the use of the derivative of the EEG might mitigate this problem and produce a more correct metric for the attenuated higher frequency oscillations.

The Teager-Kaiser Operator (TKO) was developed initially to characterize a harmonic oscillator moving at a single frequency. The idea was that the operator would characterize energy of the oscillation, thereby recognizing that oscillations usually cost more at higher frequencies. So, rather than having a power/energy parameter that exclusively depends on amplitude, the TKO employs both amplitude and frequency of the oscillation. Currently it is used in many other applications where multiple frequencies are present within a band of interest.

The analysis below outlines some of the properties and limitations of the TKO for biomedical applications and presents the derivative-alternative that might mitigate these.

##### TKO and the Harmonic Oscillator

The TKO is based on the dynamics of a harmonic oscillator. Let’s consider a mass *m* on a spring characterized by constant *k*. In a frictionless case, the position *x* (*t*) of the mass at time *t*, its velocity of movement *ẋ* (*t*) and frequency of oscillation *ω*_0_ is governed by:

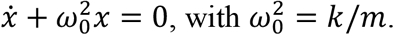

The general solution for *x*(*t*) is:

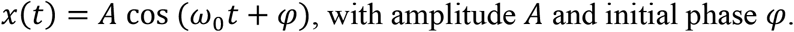

For the TKO derivation we will need the first and second derivatives of *x*(*t*), these are:

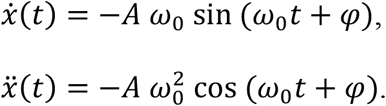

The definition of the TKO of *x*, Ψ[*x*(*t*)] is:

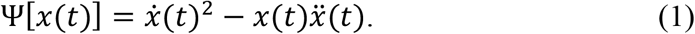

Note that the TKO is a nonlinear operator!

We now substitute the expressions we have for *x, ẋ, ẍ* in Ψ[*x*(*t*)] and we have:

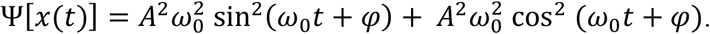

With some algebra and using sin^2^(*ω*_0_*t* + *φ*) + cos^2^(*ω*_0_*t* + *φ*) = 1, we get:

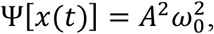

So, we find that for a harmonic oscillator, the TKO of a signal is its power *A*^2^ multiplied by its frequency squared 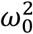, precisely what we wanted since it reflects the notion that higher frequencies require more energy to produce.

##### The TKO in Discrete Time

For sampled signals *x*(*n*) with sample number *n*, we can use the discrete version of (1). We set the sample interval equal to unity and we get:

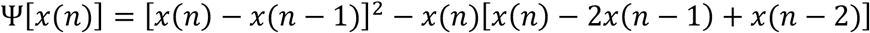

With some algebra we simplify this to *x*(*n* − 1)^2^ − *x*(*n*)*x*(*n* − 2). We now just move the index one up and get the discrete time version of the TKO:

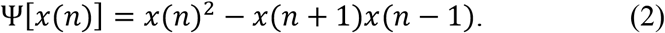

A rather simple algorithm that can provide a moving value of Ψ[*x*(*n*)].

One would say that expression is ideal for quantifying non-stationarities and variability in ongoing signals! While this is correct, the interpretation is not simply that the TKO represents frequency components multiplied by their frequencies. Due to the nonlinearity of the TKO operator, interaction across oscillations of different frequencies contribute to the result. The effect on EEG analysis of this aspect is outlined below.

##### The TKO Applied to Noise

Instead of evaluating the TKO for pure oscillations, we now consider its performance when the input is noise. To make this evaluation relevant for EEG, we consider pink noise.

We rewrite the continuous time expression (Eq. 1) in the limit form:

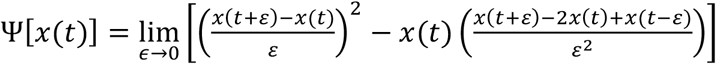, and expand the expression: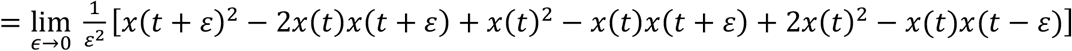.

Now we take the Expectation of Ψ[*x*(*t*)] and use:

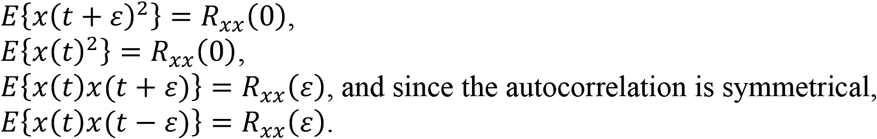

This results in:

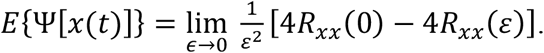

Next we apply the Wiener-Khinchin theorem to rewrite the autocorrelation as the inverse Fourier transform of its power spectrum 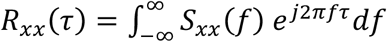. By substation we get:

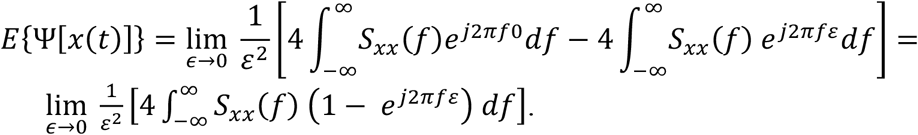

Next, we use the fact that the autocorrelation is a real even symmetric function. Hence, it’s Fourier transform consists of the (real and even symmetric cosine terms). Using this property and Euler’s equation, we can simplify the above further to:

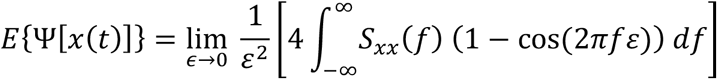

With the identity 1 − *cos*(*A*) = 2 *sin*^2^(*A*/2), we have:

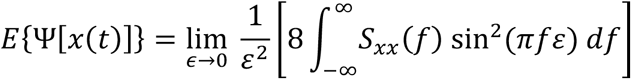

We now determine the limit and employ 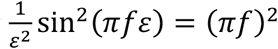. This further simplifies to:

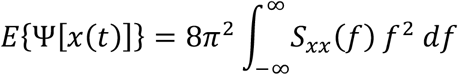

Finally we can introduce our knowledge for the bandlimited spectrum of the 1/*f* (pink) noise. The noise spectrum is a constant *C* divided by the frequency, *C*/*f*, and the bandwidth we consider is [*f*_*l*_ , *f*_*h*_]. This produces:

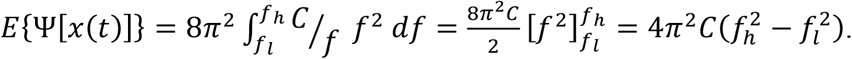

In this case we used for *C* the power of the power spectrum within the bandwidth [*f*_*l*_ , *f*_*h*_] to produce the 1/*f* spectrum of the noise.

In sum, importantly and in spite of the nonlinear aspects and ‘strange’ effects in the frequency domain, we can conclude that for characterizing the energy in the frequency band of an EEG signal, the TKO approach works quite well.

#### B. Linear Mixed-Effects Model

We fitted a linear mixed-effects model to assess the differences in gradient contrast between patient with different monogenic mutation and control and its association with age within each monogenic group. We used the following formalism:

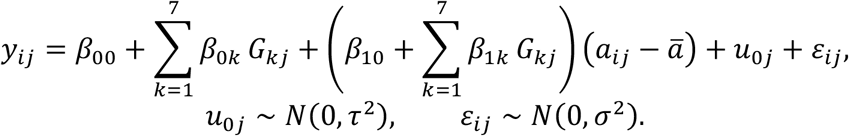

Here, *y*_*ij*_ represents the gradient contrast for recording *i* from subject *j*. Age-bin midpoints *a*_*ij*_ were centered at the mean age, *ā* = 4.87 *yr*, for all recordings. Controls is set as the reference category. *G*_*kj*_ is a categorical variable denoting the monogenic category *k* for subject *j*, where we have 7 different categories of monogenic mutation.

The coefficient *β*_0*k*_ denotes the baseline differences between the patients with monogenic category *k* and controls. The age effect on gradient contrast for control is modeled by *β*_10_. The coefficient *β*_1*k*_ represents the difference of age effect on contrast gradient between control and monogenic category *k*. Therefore, age effect for patient with monogenic mutation *k* is *β*_10_ + *β*_1*k*_.

Random intercepts and recording-level error were assumed mutually independent and normally distributed. The recordings from different subjects were assumed to be independent.

We tested the following two sets of hypotheses:

1. Th gradient contrast difference between control and patient with monogenic mutation category *k*:

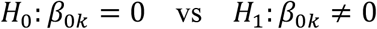
2. Age effect on gradient contrast for each group:

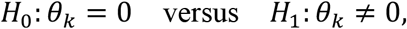

where:

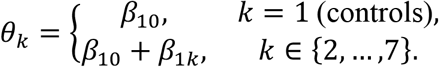

The model was fitted by restricted maximum likelihood(REML) in MATLAB. The two-sided t tests were performed using Satterthwaite degrees of freedom to account for repeated recordings. Parameter estimates were reported with 95% confidence interval. Benjamini-Hochberg correction was applied to adjust for multiple testing. An adjusted p value <0.05 is considered to be significant.

#### C. Clustered ROC Analysis

To evaluate the ability of using as gradient contrast as a biomarker to discriminate children with monogenic epilepsy from controls, we performed receiver operating characteristics (ROC) analyses using the clusteredROC() function in R (Obuchowski 1997). This method accounts for correlations among repeated EEG recordings from the same subject to compute AUC point estimate and standard deviation. Recordings were divided into two age group (1-4 and 4-10 years), and each monogenic group was compared separately with age-matched controls. The result is reported as the point estimate of AUC with the 95% confidence interval.

#### Data

EEG data are available at the sources listed in the Reference section.

All further data produced in the present study are available upon reasonable request to the authors

